# Perceived Efficacy and Acceptability of a Spatial Repellent for Malaria Control in Western Kenya: Lessons from a Qualitative Social Science Study Conducted in Conjunction with an Epidemiological Efficacy Trial

**DOI:** 10.1101/2025.09.18.25335514

**Authors:** Steven A. Harvey, Sheila Muya Ekodir, Lucy Baker, Julius Ichodo Odero, Moureen Ekisa, Jane Ikapesi, Samantha W. Tsang, Albert Casella, Kaci McCoy, Anna Passaniti, Quentin Awori, Bernard Abong’o, Brian Polo, Vincent Moshi, Sean Blaufuss, Manothri Mallikarachchi, Ashley Hudson, Nicole L. Achee, John P. Grieco, Eric Ochomo

## Abstract

**Background:** Kenyans experienced over 3 million malaria cases in 2023. Insecticide resistance and changing mosquito behavior have reduced effectiveness of insecticide-treated nets (ITNs) and indoor residual spraying (IRS). Spatial repellents (SRs) show promise for malaria control. This social science study, nested within the Advancing Evidence for Global Implementation of Spatial Repellents (AEGIS) clinical trial in Kenya, explored SR acceptability and perceived efficacy.

**Methods:** We used modified trials of improved practices (TIPs) to assess participant response to Mosquito Shield™, a transfluthrin-impregnated SR. We conducted structured observations and semi-structured interviews at 16 intervention and 14 control compounds during five visits over the 24-month clinical trial. We remained blinded to study assignment throughout data collection then analyzed data by study arm after unblinding.

**Results:** Both intervention and control participants reported reduced mosquito activity inside homes after initial installation. Over time, most intervention participants reported continued efficacy. Controls, who received identical-looking placebo products, reported decreased efficacy. In both arms, participants mentioned discontinuing ITN use. Closely tied to perceived efficacy, acceptability was also influenced by installation method, number of units required, replacement interval, and perceived community opinion.

**Discussion/Conclusion:** SRs seem likely to be accepted in this setting when perceived as effective and installed satisfactorily. National programs considering SR distribution should anticipate decreased ITN use and emphasize that SRs are meant to complement, not replace, existing interventions. SR-related communication should stress proper product use to maximize efficacy. Decreased expenses for products like mosquito coils might incentivize adoption. This study illustrates the value of social science research parallel to an efficacy trial.

## Background

Malaria remains a public health priority in Kenya, with an estimated 3.3 million cases in 2023.[1] With one of the country’s highest prevalence rates, Kenya’s Lake Region is a focus for vector control including insecticide-treated nets (ITNs), indoor residual spraying (IRS), and larval source management (LSM).[2] While these interventions have reduced the disease burden, residual malaria transmission remains a challenge.[2–4] ITNs are only effective when used during peak anopheline mosquito biting hours. Insecticide resistance and early morning biting when many people are awake and outside their ITNs further limit protective efficacy.[5–7] Additional tools are urgently needed to fill protection gaps.

Spatial repellents (SRs), designed to prevent indoor mosquito-borne pathogen transmission, show promise.[8] *Advancing Evidence for the Global Implementation of Spatial Repellents* (AEGIS) conducted a 24-month cluster-randomized controlled trial (cRCT) of Mosquito Shield™, a transfluthrin-impregnated SR, against malaria in Busia County, Kenya.[9, 10] The WHO Vector Control Advisory Group (VCAG) confirmed that the Kenya cRCT results “demonstrated the public health value of the spatial repellent for the prevention of malaria, above that provided by LLINs alone…”[11].

However, moving from trial-based efficacy to real-world effectiveness requires that household users see SRs as acceptable and safe. Understanding factors likely to influence acceptance – including perceived efficacy, comparison to existing methods, and ease of use – is crucial.[12] Failure to confirm acceptability may result in low uptake, discontinued use, or outright rejection. [13–16] Further, community input may spur modifications that enhance product acceptability to future users.

This paper presents social science findings about facilitators and barriers to SR adoption and sustained use. Programmatic recommendations on procurement, promotion, and distribution are based upon cRCT participant feedback.

## Methods

Study site characteristics and details of the parent cRCT to which this study was linked are described elsewhere.[9, 17] Briefly, Busia County is located in Western Kenya near Lake Victoria and the Ugandan border. The region experiences year-round malaria transmission with peaks during the long and short rains. Villages consist of multi-generational family compounds typically including a main house or hut occupied by the family patriarch or matriarch plus additional structures for grown children and their spouses and children. Some compounds contain structures to cook, store grain, and house animals. The cRCT enrolled 3781 compounds from 58 clusters, 1896 randomly assigned to receive transfluthrin-impregnated SRs (intervention) and 1885 an identical-looking placebo (control).[17] Researchers, study team members, and participants were all blinded to study assignment.

### Mosquito Shield™ spatial repellent characteristics

Mosquito Shield™ is a clear plastic sheet, measuring 8.5” x 12.5” installed, treated with a standard quantity of transfluthrin which it passively emanates throughout its 28-day lifespan. Manufacturer specifications stipulate installation of two units per 9 m^2^ of interior space, at 2-3 m above ground. Products were hung on mud, cement and brick walls using double-sided tape or metal hooks (Figure 1). In structures with fabric-covered walls, safety pins were used.

**Figure 1:**
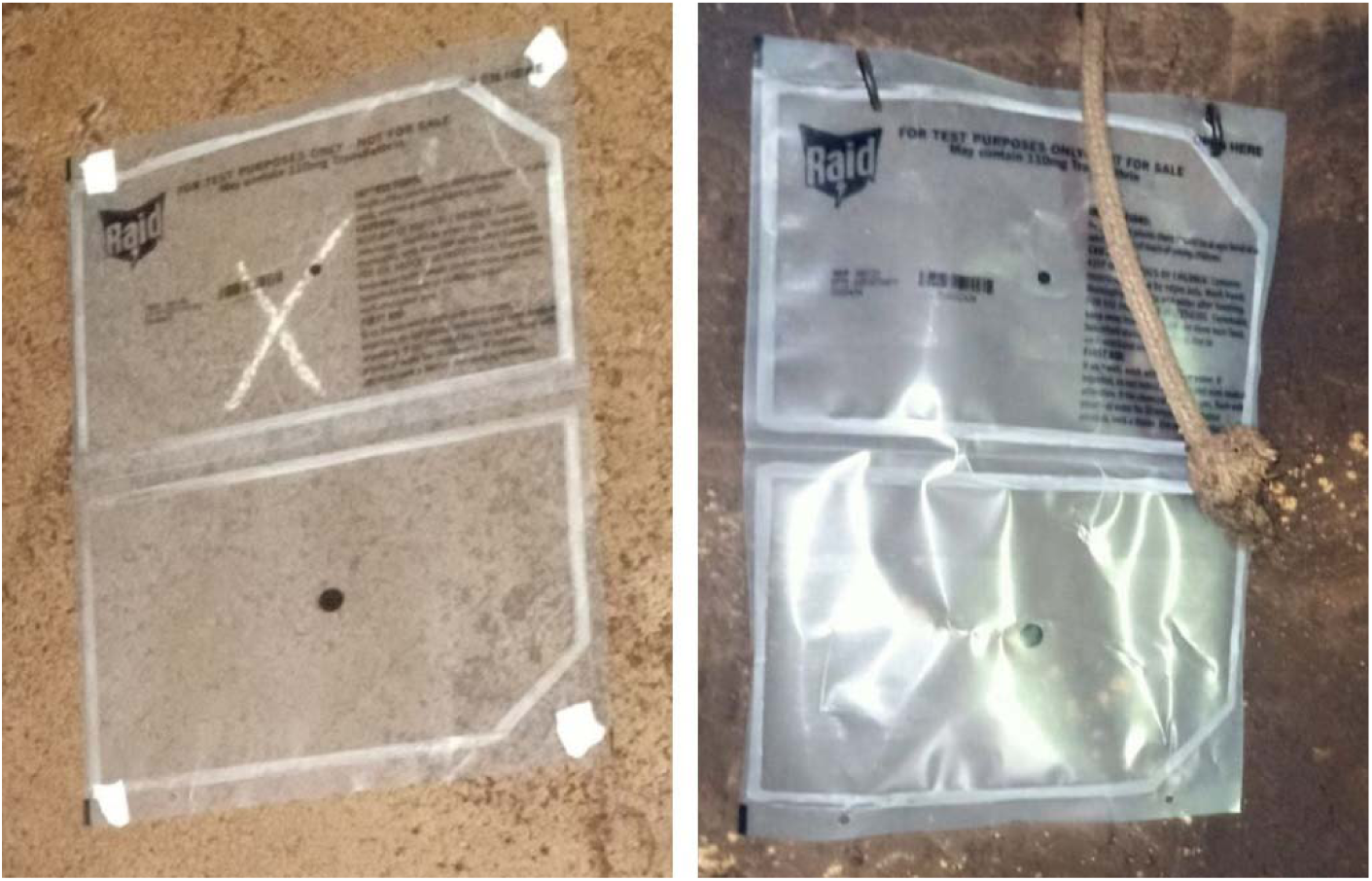
Mosquito Shield^TM^ installed inside houses with tape (l) or hooks (r)

### Data collection method

#### Trials of improved practices (TIPs)

To assess perceived efficacy and acceptability and solicit input on improving product characteristics, we employed modified trials of improved practices (TIPs). TIPs are a participatory method designed to seek community input on the feasibility and acceptability of a proposed health behavior change intervention prior to introduction. Researchers typically recruit 15-20 participants to pilot-test the proposed intervention over several cycles and modify it into something capable of achieving the public health objective and feasible for the target population to implement.[18, 19] For example, Leontsini, et al., used TIPs to test and modify improved water storage practices to eliminate *Aedes aegypti* breeding sites and reduce dengue transmission in the Dominican Republic.[20] TIPs studies usually recruit participants purposively because participation requires extended commitment to the study and repeated contact with the study team, a burden not all households are willing to accept. Participants must also be willing to set aside social courtesy bias when reporting problems.

#### Modified TIPs

We label our method “modified TIPs” because we sought input on a product (Mosquito Shield™) not a behavior. Questions included where and how to install the SRs, information participants wanted about them, aesthetic preferences (e.g., shape, color, graphics, size, smell) plus any additional issues that surfaced during the trial.

A study team member made 5 post-installation visits to each participating compound: at one week, then two, six, 12 and 18 months.

Visits began with a structured observation noting whether each installed SR was hanging as intended or had become partially dislodged from the wall, fallen, was covered by or touching some other object, or appeared damaged, handled or otherwise modified. The team observed two structures per compound: the main structure occupied by the family head and either a separate structure in which children slept or the kitchen if the compound had no children or children slept in the main structure. Observers collected data on a tablet computer using a 26-item questionnaire programmed in CommCare^©^.[21]

Study team members then conducted a semi-structured qualitative in-depth interview (IDI) with a resident adult asking about the family’s experience with the SRs and any anomalies noted during the observation. Whenever possible, study team members visited the same compounds and interviewed the same participants during each cycle. Interviews were digitally recorded in Kiswahili or Ateso based on participant preference, transcribed in the original language, and translated into English for analysis.

#### Sampling

To protect the cRCT’s double-blinded design, we used two-stage sampling to select 30 compounds. First, the unblinded data safety monitoring board (DSMB) statistician drew a random sample of 12 clusters, six intervention and six control. We then purposively selected 2-3 compounds per cluster. Community health volunteers and the study team helped identify participants who met TIPs inclusion criteria. We remained blinded to assignment until after completion of data collection and cRCT efficacy analysis.

### Data Analysis

For qualitative data, study team members developed an initial codebook based upon the study’s research questions (deductive) and a preliminary data review (inductive). To reach consensus among coders, we double-coded 15 randomly selected transcripts, meeting repeatedly to resolve interpretive differences. We also wrote and shared analytical memos tracking our evolving understanding of codes and themes.

We used ATLAS.ti 24 to assist with coding and data management.[22] We then exported coding results to Microsoft Excel creating chronological matrices by theme: perceived efficacy, side effects, suggested improvements, and comparison to other mosquito control products. We charted each theme onto a separate worksheet with compounds (1-30) in rows and visits (1-5) in columns. Quotations corresponding to a theme and compound were pasted into cells by visit. This yielded a matrix in which we could trace the evolution of any theme for any compound chronologically from left to right across a single row.

Coding and initial matrix building took place with still-blinded data. After unblinding, we stratified by study arm to produce comparative interpretations. Weekly meetings to discuss interviews, coding, and thematic analysis helped identify and refine emerging themes. Meetings took place in person in Busia until December 2022, then continued virtually between Kenya- and US-based team members.

For quantitative data, the study team determined mean, median, or proportion as appropriate and calculated p-values where possible and relevant.

### Research Ethics

The study received ethics approval from the Kenya Medical Research Institute (KEMRI) Scientific and Ethics Review Unit, the Johns Hopkins Bloomberg School of Public Health (BSPH) Institutional Review Board, and the World Health Organization (WHO) Ethics Review Committee. All participants provided written informed consent in Ateso, Kiswahili, or English as they preferred. Non-literate participants consented in the presence of a literate witness of their choice.

## Results

The team enrolled 30 compounds, 16 intervention and 14 control. One intervention compound exited the study after TIPs visit 2 due to concerns about side effects, leaving 15 intervention compounds and 14 controls. Observation data for the compound lost to follow-up are included through visit 2. In addition, one control participant was unavailable in round 3 while one intervention participant was unavailable in rounds 2 and 4, leaving an overall sample of 143 interviews, 75 intervention and 68 control (Table 1).

**Table 1:**
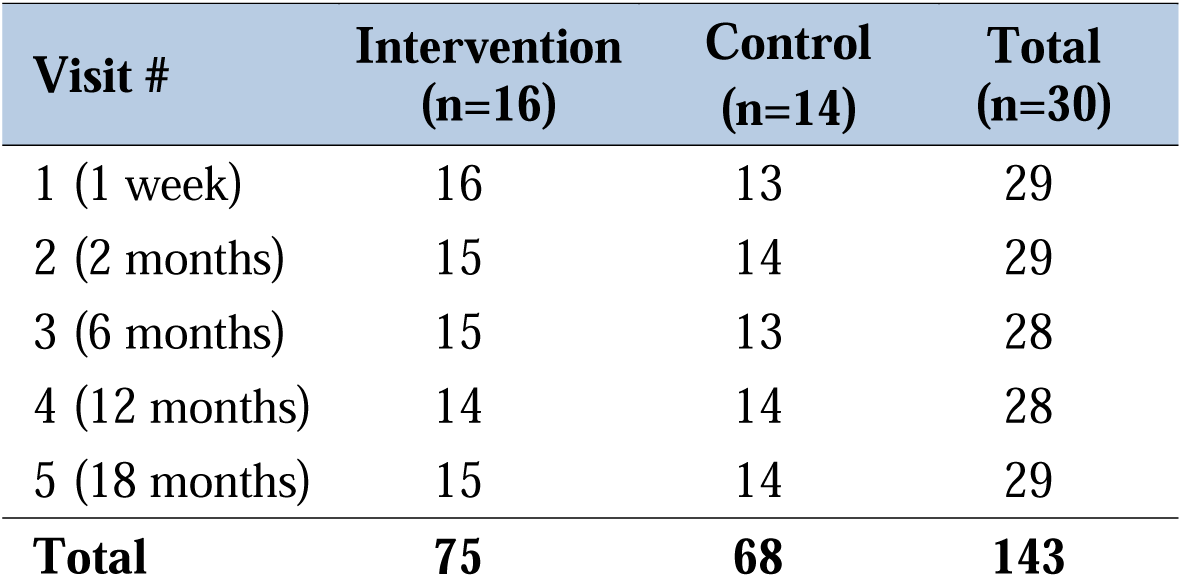
In-depth interviews by TIPs visit (1-5)

| **Visit #** | **Intervention**<br>**(n=16)** | **Control**<br>**(n=14)** | **Total**<br>**(n=30)** |
| --- | --- | --- | --- |
| 1 (1 week) | 16 | 13 | 29 |
| 2 (2 months) | 15 | 14 | 29 |
| 3 (6 months) | 15 | 13 | 28 |
| 4 (12 months) | 14 | 14 | 28 |
| 5 (18 months) | 15 | 14 | 29 |
| **Total** | **75** | **68** | **143** |

### Observation Results

There were no statistically significant differences between intervention and control compounds in mean number of rooms observed, SRs installed, or interviewee age or sex (Table 2).

**Table 2:** Characteristics of TIPs compounds by study arm assignment.

| **Compound** | **Intervention (n=16)** | **Control (n=14)** | **p-value** |
| --- | --- | --- | --- |
| Mean rooms in structure | 5.3 | 4.9 | 0.67 |
| Mean products installed | 17.3 | 16 | 0.61 |
| Interviewee age (mean) | 48.1 | 52.1 | 0.46 |
| Interviewee sex (% female) | 50 | 50 | - |

Among observed intervention and control structures combined, the total number of observed rooms per round varied slightly from a high of 140 in round 1 to a low of 135 in round 5. Variation in the number of rooms in a given compound across time was due either to a room being inaccessible during a particular visit or to housing modifications made during the trial. Intervention compounds had a higher number of rooms observed (mean 72.6, SD 1.52) than control compounds (mean 65.0, SD 1.22, p<0.001). However, the number of SRs required per room did not differ significantly between intervention (mean 3.2) and control (mean 3.3, p=0.56).

A key TIPs question was whether participants might remove, lose, alter, or otherwise manipulate SRs. As shown in Table 3, removal, loss, and manipulation were rare in both trial arms. This remained consistent and stable throughout all five rounds. Round 2 showed slightly more handling among control compounds (0.2) than intervention (0.1, p=0.05); round 5 showed the opposite (0.4 intervention, 0.0 control, p=0.02). Though statistically significant in both cases, the actual number of products affected did not seem programmatically meaningful. The situation is similar for SRs observed to be somewhat worn in rounds 2 and 5.

**Table 3:** Status of Mosquito Shield^TM^ products expected (2 units per 9m^2^) and observed by TIPs observation round^†^.

|  | Round 1 (1 week) |  |  | Round 2 (2 mos) |  |  | Round 3 (6 mos) |  |  | Round 4 (12 mos) |  |  | Round 5 (18 mos) |  |  |
| --- | --- | --- | --- | --- | --- | --- | --- | --- | --- | --- | --- | --- | --- | --- | --- |
|  | Int | Con | p | Int | Con | p | Int | Con | p | Int | Con | p | Int | Con | p |
| SR presence or absence - missingness |  |  |  |  |  |  |  |  |  |  |  |  |  |  |  |
| # rooms observed | 73 | 67 | □ | 73 | 65 | □ | 74 | 65 | □ | 73 | 64 | □ | 70 | 64 | □ |
| # SRs required in room | 3.2 | 3.3 | 0.65 | 3.2 | 3.3 | 0.81 | 3.2 | 3.3 | 0.76 | 3.2 | 3.3 | 0.86 | 3.2 | 3.2 | 0.91 |
| # SRs actually installed in room | 3.2 | 3.3 | 0.83 | 3.2 | 3.2 | 0.96 | 3.3 | 3.3 | 0.98 | 3.2 | 3.3 | 0.81 | 3.2 | 3.2 | 0.99 |
| # SRs hanging on wall | 3.2 | 3.3 | 0.93 | 3.2 | 3.2 | 0.94 | 3.2 | 3.1 | 0.75 | 3.2 | 3.2 | 0.82 | 2.9 | 3.2 | 0.36 |
| # SRs visible but not hanging on wall | 0 | 0 | 0.3 | 0 | 0 | 0.6 | 0.1 | 0.1 | 0.39 | 0 | 0 | 0.93 | 0.2 | 0.0 | 0.14 |
| # SRs not visible and not hanging on wall | 0 | 0 | 0.95 | 0 | 0 | □ | 0 | 0 | 0.29 | 0 | 0 | □ | 0.0 | 0.0 | 0.45 |
| SR condition when observed |  |  |  |  |  |  |  |  |  |  |  |  |  |  |  |
| # SRs not handled / disturbed since installation | 3.2 | 3.2 | 0.81 | 3.2 | 3 | 0.55 | 3.1 | 3.1 | 0.87 | 3 | 3.1 | 0.51 | 2.8 | 3.1 | 0.17 |
| # SRs handled / disturbed since installation | 0.1 | 0.1 | 0.68 | 0.1 | 0.2 | **0.05\*** | 0.1 | 0.2 | 0.59 | 0.2 | 0.1 | 0.24 | 0.4 | 0.0 | **0.02\*** |
| # SRs physically intact and clean | 3 | 3.2 | 0.61 | 3.1 | 2.9 | 0.42 | 3 | 3 | 0.99 | 2.7 | 3 | 0.23 | 2.7 | 3.1 | 0.15 |
| # SRs somewhat worn | 0.2 | 0.1 | 0.48 | 0.1 | 0.3 | 0.07 | 0.2 | 0.2 | 0.99 | 0.5 | 0.2 | 0.1 | 0.5 | 0.1 | **0.03\*** |
| # SRs very worn | 0 | 0 | □ | 0 | 0 | □ | 0 | 0 | □ | 0 | 0 | □ | 0.0 | 0.0 | □ |
| Installation method |  |  |  |  |  |  |  |  |  |  |  |  |  |  |  |
| # SRs installed with tape (4 tapes per SR) | 1.6 | 1.4 | 0.62 | 1.6 | 1.4 | 0.72 | 1.6 | 1.5 | 0.56 | 0.4 | 0.1 | 0.06 | 0.0 | 0.1 | 0.3 |
| % SR tape remaining | 69% | 93% | **<0.001\*** | 73% | 85% | **0.04\*** | 65% | 51% | **0.01\*** | 49% | 53% | 0.65 | □ | 25% | □ |
| % SR tape missing | 8% | 0% | **0.03\*** | 26% | 14% | **0.04\*** | 21% | 45% | **<0.001\*** | 34% | 47% | 0.55 | □ | 75% | □ |
| # SRs installed with hooks (2 hooks/SR) | 1.6 | 1.7 | 0.79 | 1.6 | 1.8 | 0.64 | 1.6 | 1.8 | 0.56 | 2.8 | 3.1 | 0.27 | 3.1 | 3.0 | 0.46 |
| % SR hooks remaining | 102% | 100% | 0.44 | 101% | 100% | 0.47 | 98% | 98% | 0.7 | 96% | 98% | 0.28 | 94% | 98% | 0.23 |
| % SR hooks missing | 0% | 0% | □ | 0% | 0% | 0.23 | 3% | 2% | 0.73 | 4% | 1% | 0.16 | 4% | 3% | 0.41 |

<sup>†</sup> Numbers rounded to nearest two decimal places; “mos” = months, “Int” = intervention, “Con” = control; observation round refers to time from first product installation in the home
\* $p \leq 0.05$

While most SRs remained in place from installation to replacement, observers frequently found SRs partially or fully covered by posters, other wall-hangings, or clothes. Participants also often used hooks to hang additional objects alongside or atop the SR (Figures 2-3). Observers frequently found that SRs installed in kitchens were covered with soot from cooking fires (Figure 4). In other rooms, SRs often became coated with dust.

**Figure 2:**
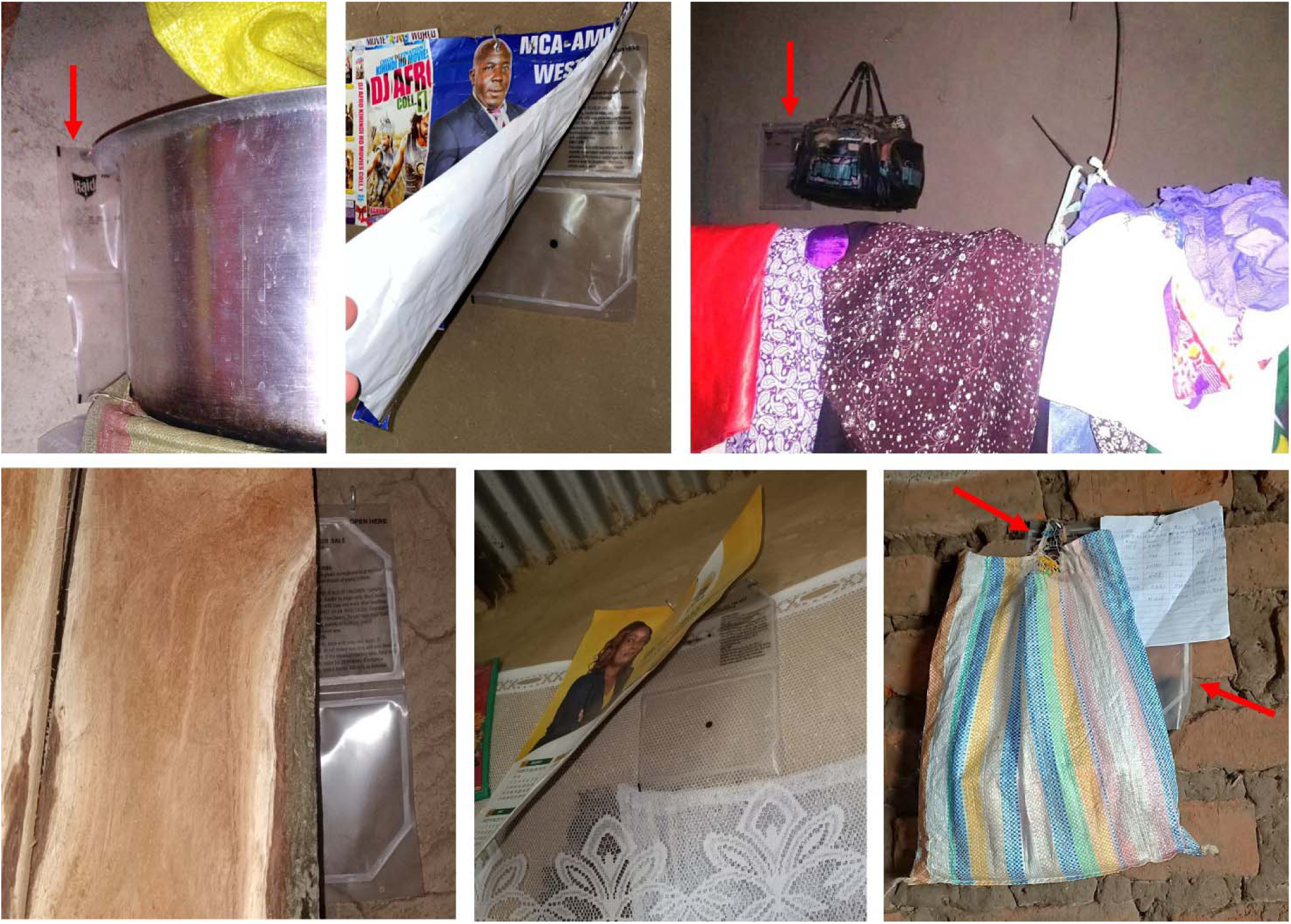
Mosquito Shield^TM^ partially or fully covered by other household objects.

**Figure 3:**
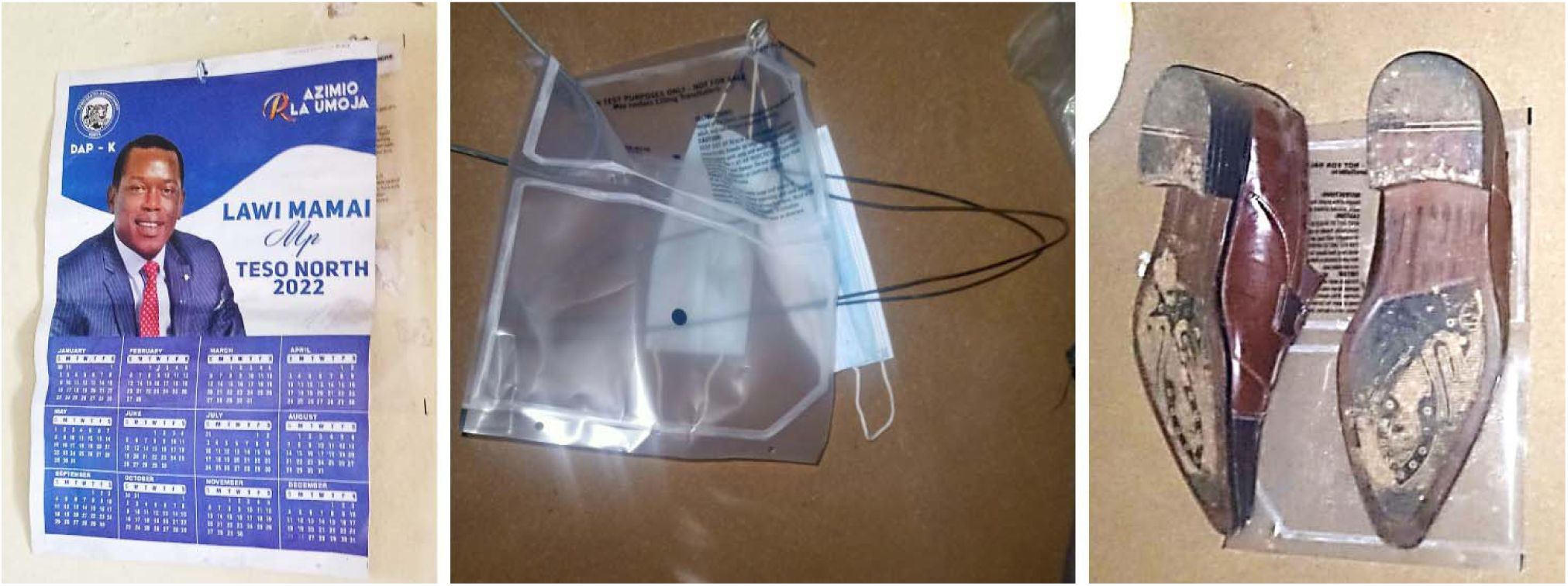
Study participants used hooks from Mosquito Shield^TM^ installation to hang other household objects.

**Figure 3:**
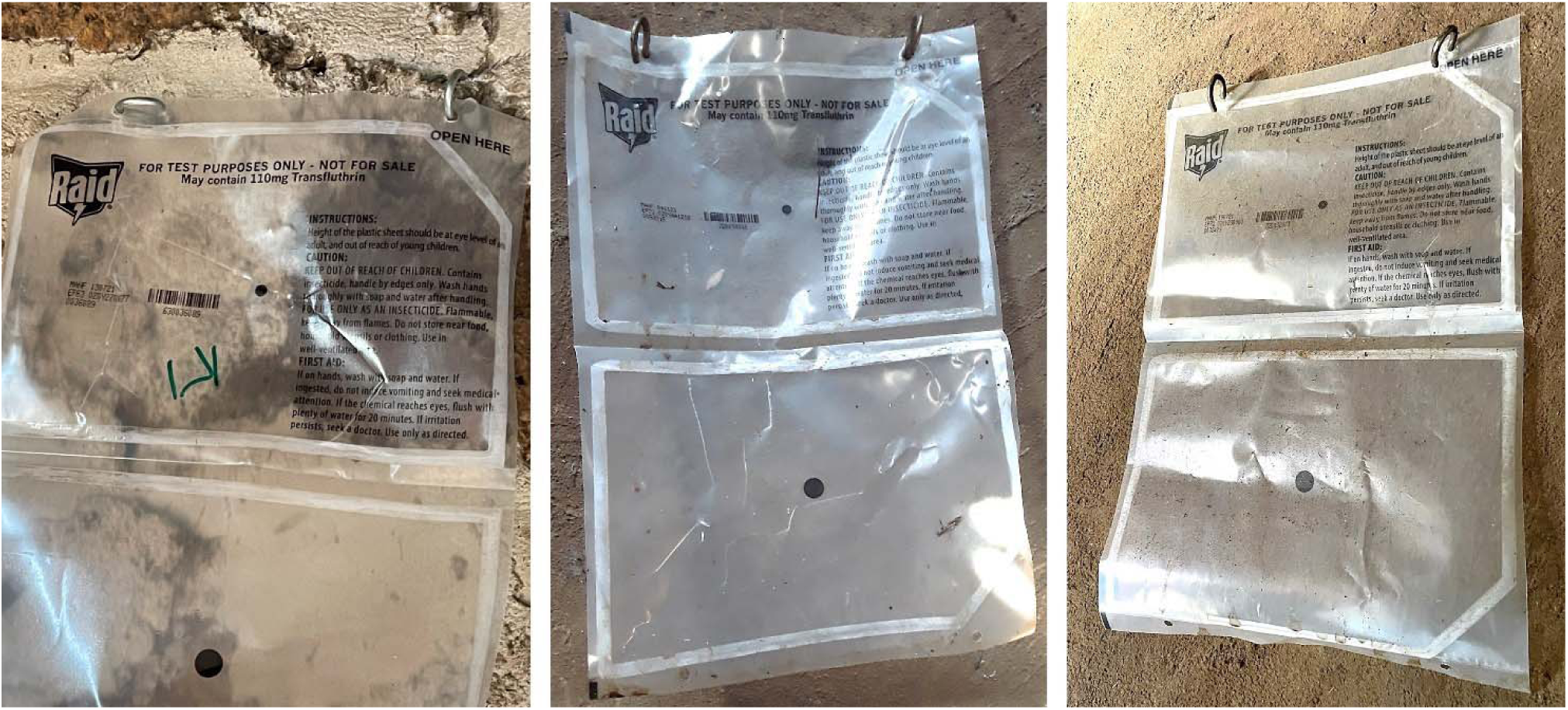
Hooks rotated in different directions to prevent product from blowing off the wall.

**Figure 4:**
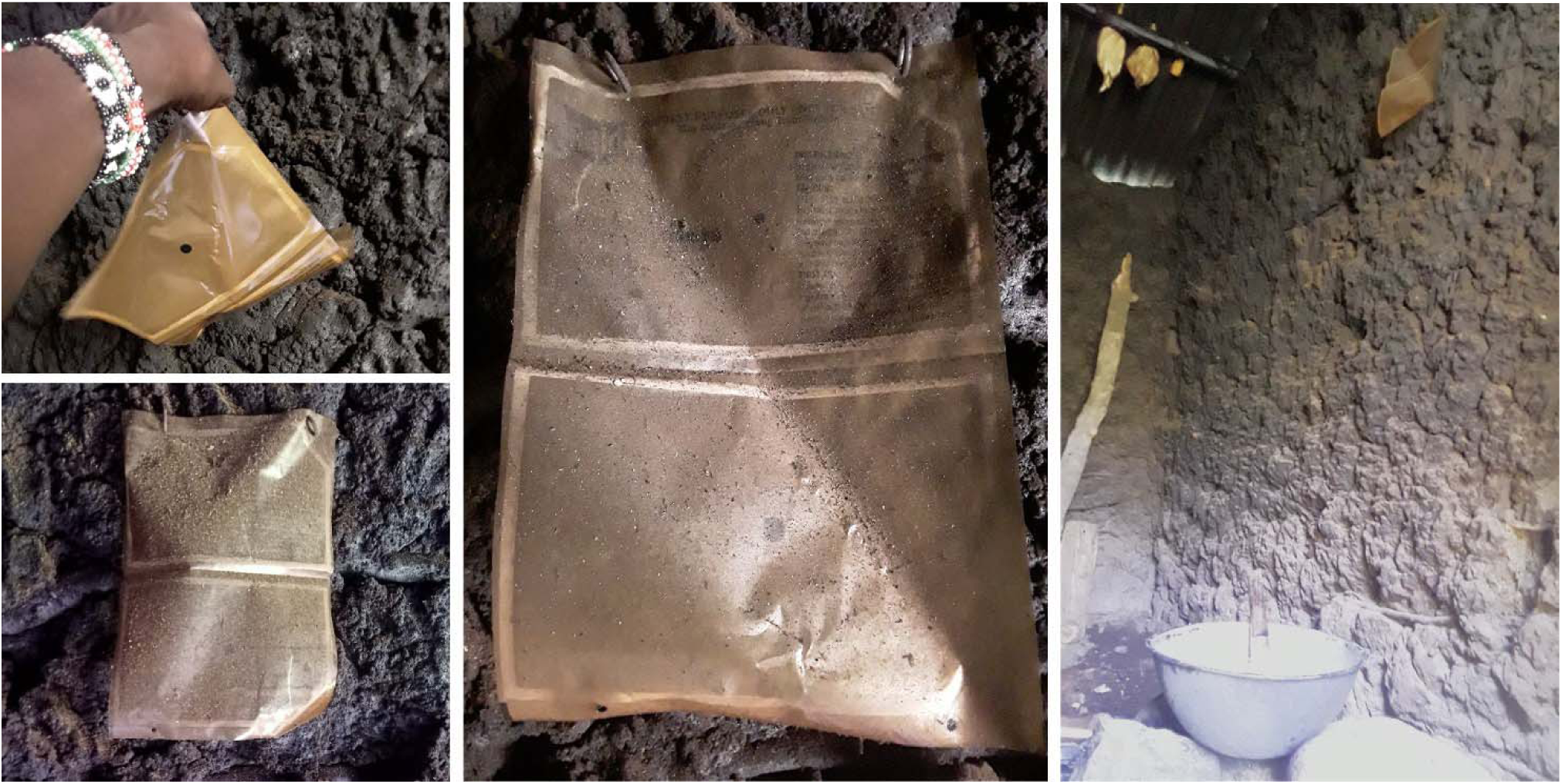
Mosquito Shield^TM^ showing soot from cooking fire, photos taken in same kitchen but at different times and from different angles.

### Perceived Efficacy

In round 1 visits, one week after initial installation, most intervention and control participants stated that they perceived the SR to be effective because they had seen fewer mosquitoes. From round 2 onward, most intervention participants reported that mosquito density began rising again near the end of every 28-day replacement period. Most controls reported no decline, or a brief decline followed by a rapid return to normal. This was not universal, however: some intervention participants in some rounds reported that SRs were not effective while some controls stated that they were.

Participants described SR efficacy principally in terms of mosquito volume or density and vigorousness of activity (flight speed, buzzing, biting aggressiveness).

> Since you installed these products… the mosquito population has completely reduced. I can say by 99% maybe when you are going to sleep is when you will hear one making a sound. But those that are found you will see them drunk, flying slowly, and you will even see them fall down by themselves. (Male, 30’s, intervention, round 1)

A smaller number described efficacy in terms of reduced malaria incidence:

> Now even the state of malaria too, there’s this girl of mine that is in grade six, she really used to like falling sick. After just like one and a half months like this… malaria gets to her mostly. But we see it has come down since that time This year since January we’ve only treated once. (Male, 40’s, intervention, round 3)

Table 4 shows illustrative comments about efficacy across the five rounds of TIPs. Some intervention and control participants also mentioned a reduction in insects such as cockroaches, wasps, and bedbugs. However, some said they saw no change, and others reported a decrease in initial rounds followed by a return to previous levels.

**Table 4:** Illustrative comments about perceived SR efficacy across the 5 rounds of TIPs.

| Study arm | Round 1 (1 week) | Round 2 (2 months) | Round 3 (6 months) | Round 4 (12 months) | Round 5 (18 months) |
| --- | --- | --- | --- | --- | --- |
| Intervention | These drugs really chase mosquitoes away. Though other mosquitoes are blown by the wind to the house but those that used to stay inside the house are no longer there. Even those that are blown into the house you just hear them make noise a little, then shortly they disappear or die. | In the first month, I observed few mosquitoes, there were many but have seen only like 3 so far...<br>That *dawa* [insecticide] is like against them: Immediately they sense the product, they flee away. | These things are doing a very good job. So, me, I was now thinking since it is a project and I am in the research, how long will it take and in what state will you leave people? Because once you remove these things those mosquitoes will still just come back. | If I compare to the past, the population of mosquitoes was high and I see that they are decreasing. With the dry season which usually has many mosquitoes, especially ...when the rain is not there, mosquitoes are usually very many. So, I have seen here... this project, only once in a while I hear one buzzing. | I don't know where they came from, but all that time I haven't seen so many of them. Even if you sit in the living room like this, it's not easy for mosquitoes to bite you, mmmh. Even when you sleep it's not a lot, mmmh. |
| Control | I see those medicines [the SR] are working well. Since those medicines were placed, I have not seen what we call a mosquito. And I thank you for bringing the medicine. | What I have seen, they [the SRs] have managed to reduce mosquitoes. Their paining is not much at all [they are not bothering us much at all]. | In the past, the mosquitoes were biting, you see when you are seated like this, you felt them biting you. But at the moment since they installed these [SRs], I see it is reducing. | Some are saying that those [SRs] help to chase away mosquitoes, but others say that they have brought these things, and mosquitoes have now increased. | There was a difference in the past when you first put these things, but now, I see it is like they [the mosquitoes] have increased and they have become aggressive, *eeeh* [yes]. |

| Study arm | Round 1 (1 week) | Round 2 (2 months) | Round 3 (6 months) | Round 4 (12 months) | Round 5 (18 months) |
| --- | --- | --- | --- | --- | --- |
| Intervention | ...in the past one week I have noticed reduction in mosquitoes. My house had a lot of mosquitoes. When you were seated at night, they used to bite the legs, bite the hands. But since they installed these products, I have noticed mosquitoes have reduced in the house. | In these three months, I still don't see many mosquitoes. Mosquitoes have reduced I have seen just a few and they are not like they used to be in the past when these products were not there. | I see these products are good, they are working. Because just as I said previously, if you compare mosquitoes outside... and here in the house, I see mosquitoes have reduced in the house as compared to out there because I have [SRs] inside... | So when these people came to install [SRs], they told me that some have *dawa* and others don't have *dawa*. So when you are lucky and get the ones that have *dawa*, that repels mosquitoes. But when you get those that don't have *dawa* mosquitoes will still be there. | *Eeeh* now, when they put the one with the drug, mosquito activities will reduce. And when the ones without the drug are put, mosquito activities, will just continue I mean they will be high, mmmh |
| Control | What I can tell you, I cannot lie: I have seen bad things since [the SRs] were placed. Like yesterday my neighbor said that in [her] place it is as if those plastics went and brought mosquitoes... 'These people went and invited the mosquitoes where they were.' Then I said, 'Aaah! ...come and see my place.' When she came, she said, 'It is the same plastics.' | When those things are placed, when they are still new like that, you don't see any mosquitoes. After a short time like how many, when its strength gets finished, you again start seeing mosquitoes. | ...these ones are tired, they are not working. *Eeeh!* These have become tired. Because I was told [by neighbors] for sure, if... you... come to ask questions..., tell them that the plastics have refused to work. Mosquitoes have increased. *Eeeh!* Everyone is complaining, mosquitoes have increased there is no benefit. | (Comparing SRs to an ITN):<br>A bed net, when you have gone under it, there is no place will pass to bite you [laughs]. And now these [SRs], if you are in the sitting room here will the mosquito not bite you? It will bite you! ...when you fan on this side, it again flies and passes on the other side... | During rainy season, mosquitoes are many, but now since the rains have begun skipping, they are few. Then, on these papers [SRs], some have *Ekeya* [active ingredient], others do not have *ekeya*. Like the ones that were replaced yesterday, the ones [CHVs] who, removed, they had nothing at all. Mosquitoes were a lot in the house. |

### Perceived Acceptability

Participant comments about acceptability were closely aligned with perceived efficacy. At the 1-week visit, both intervention and control participants spoke positively about the SRs and expressed optimism about their effectiveness. Controls continued to express hope for the product at the 2-month and 6-month visit but also reported growing skepticism about effectiveness. Controls commented that the SRs seemed to work more effectively for some families or in some rounds and not others, leading to doubts about reliability. Skepticism grew over time, with some stating that community members were calling the SRs useless or saying they attracted mosquitoes instead of repelling them.

Intervention participants, meanwhile, reported positive views of SRs through all five TIPs rounds. In addition to perceived efficacy, factors favoring acceptability included a reportedly significant reduction in other insects like wasps and cockroaches, protection outside sleeping hours, and a relatively unobtrusive presence:

> It makes me happy when they [SRs] are stuck on the wall. They are not something that soils the house, they are not things of dust [i.e., they don’t create a mess or attract dust], it is just something that is sticking on the side. Why now should I be sad? If it were something that fans dust, and you see it brings something bad ahead, maybe bad smell, is when you can say, ‘Aaah, these people have also brought things with another smell’ But like those ones [SRs], what can you say? Something that you cannot see smelling, something you do not fan, you do not see. Again, if someone hasn’t looked at your wall, they cannot even notice. They cannot even know… I will tell them; those things are good. Me who has accepted those things, the dirt [bother] of those things that bite the body [i.e., mosquitoes] has ended. I slept continuously [without interruption]. I do not see again something biting me or maybe flying, no …in my house, no. That is why I have embraced those things. (Male, 70’s, intervention, round 3)Many expressed a desire for SRs to be expanded to neighboring communities and additional structures such as toilets and animal housing, within their compounds.

A potential barrier to acceptability was concern about harm to children. One participant in her 60’s mentioned her grandchildren picking up SRs that had fallen off the wall:

> The way they told us that children should not touch,” she added, “we were worried. My grandchildren are very cheeky. That is why I said they should remove these things [SRs]… The products would scatter all over the kitchen whenever they fell, and the children would quickly pick them up. That is why I saw there was no need. (Female, 60’s, intervention, round 4)Of the 3781 cRCT participants, this participant was one of 11 (5 intervention and 6 control) to withdraw from the study.[17]

### Side effects

Participants commented on SR side effects in 114 of the 143 interviews (56 intervention, 58 control). Table 5 shows a distribution of those comments by study assignment arm.

**Table 5:** Did participant report perceived SR-related side effects in interviews mentioning side effects? # (%)

| **Study arm** | **No** | **Unsure** | **Yes** | **Total** |
| --- | --- | --- | --- | --- |
| Intervention | 41 (73) | 9 (16) | 6 (11) | 56 (100) |
| Control | 37 (64) | 10 (17) | 11 (19) | 58 (100) |
| **Total** | **78 (68)** | **19 (17)** | **17 (15)** | **114 (100)** |

The great majority in both groups reported no side effects in humans. One intervention participant noted that while the SRs had a smell similar to an ITN, it was less bothersome:

> It has been put in a way it does not irritate you like the one that is in [an ITN]… [In a] mosquito net, it irritates the nose a lot. That is why I am telling you, even for an asthmatic person, I have never heard of a complaint. Especially my mother, since this product was installed in her house, I have never heard her complain that it is affecting her breathing. (Male, 30’s, intervention, round 2)

Earlier in the same interview, this participant reported that his family had stopped using mosquito coils after receiving SRs because his wife was asthmatic and the coils caused her breathing problems. A different asthmatic participant told the interviewer that he feared he would have breathing problems when the SRs were installed but had not. He said he detected no odor from the SRs and preferred an odorless product such as this because it was less likely to aggravate his asthma.

Some described conversations with neighbors concerned about potential illnesses attributable to the SRs:

> Those who don’t have in their houses do ask me, ‘you who have those ikee [drugs, i.e., SRs] in your houses, how do you feel?’ And I say, ‘we who have those ikee, we have not seen any harm from mosquitoes, mosquitoes are not many.’

> ‘And of the reports that it causes peeling of the skin, and it irritates the eyes?’ And I said, ‘I have not seen.’

> ‘And if you touch by mistake? What signs do you see in your hands?’ And I said, ‘Aaah! [No] I have mine, which is hanging on my bags. I do touch every time, but I have not seen any signs.’

> *[A neighbor responded] that ‘no, they are saying that paper, it irritates the eyes, that if you touch, I don’t know what it does.’ And I said, ‘no.’ (Male, 50’s, intervention, round 2)*Most participants reported no side effects on dogs, cats, chickens, or other farm animals. Seven intervention and 11 control participants reported being unsure whether human or animal illnesses were due to the SRs or to natural causes. Six intervention and 11 controls reported allergic reactions such as eye or nose irritation or coughing:

> There was [a] time my brother’s child, who stays here, I don’t what he was looking for up there, so when he got closer to the Ikee, he started sneezing. Eeeh. And I told him it is not allowed to be close to this Ikee. (Male, 40’s, intervention, round 2)“The children also started sneezing,” reported a different participant, “and I had to take them for medication. Sneezing has become a lot; headache has become a lot.” (Male, 40’s, intervention, round 2). Asked if the children had a similar reaction when the SRs were first installed or after the first replacement, the interviewee said it had happened only after the second replacement. In round 3, one intervention participant reported a smell that caused burning and itching. However, he reported no side effects by the final round.

### Effect on ITN and Other Mosquito Control Use

The Kenya National Malaria Control Program distributed new long-lasting ITNs in Busia County just prior to trial launch. Study staff advised all participants to continue using ITNs since SR efficacy was still unconfirmed and half participants would be receiving a placebo. Nevertheless, over half participants in both arms reported abandoning ITN use at some point during the study – usually during the first two rounds.

Most later reported resuming net use, but often described it as occasional, adding that they would decide based on their perception of SR effectiveness. If there were few mosquitoes, they would forgo use; if mosquitoes increased, they would resume. Many reported resuming after renewed CHV or study staff encouragement. Some stated that they resumed out of habit or for additional protection rather than because they considered it necessary. Households that never resumed use attributed this to perceived SR efficacy.

> Since [the mosquitoes] disappeared, I have not seen any problem. I just sleep without [a net]… when they install, you will not hear them [mosquitoes] buzzing again at all… You just sleep without [a net] up to the morning. (Female, 70’s, intervention, round 5)

More intervention than control participants reported discontinuing use of other mosquito control methods. As one explained:

> …what has made us stop using these mosquito coils and mat is because of these products [SRs]. Because we have seen there are no more mosquitoes, now you will burn to do what [for what purpose]? (Male, 30’s, Intervention, round 1).Throughout the trial, however, participants in both arms mentioned continued use of other mosquito control methods outdoors and in locations lacking SRs. They also mentioned continuing to cut grass and remove standing water from around their compounds, activities they described as both a way to prevent mosquito breeding and a routine household duty.

### Community-recommended improvements

In early rounds, most participants in both groups said they had no specific preference about SR size, shape, color, or smell. However, requests for more powerful or longer lasting SRs became more common during later rounds.

> Ekeya [active ingredient] needs to be added because it is as if it is easy for its ekeya to get finished within a short time because the day it was placed that ekeya was smelling but I see no. I see that ekeya is [weak], its strength is little. They should add another one that has more strength. (Male, 60’s, control, round 2)Suggested replacement intervals ranged from two months to five years. Reasons included establishing a routine, maintaining privacy, and ensuring someone was home when CHVs came with replacements. Two participants mentioned price as justification for longer replacement intervals: if SRs were expensive, it would be difficult to buy them more than every 3-4 months.

Most control group participants seemed to assume that their products had insufficient *dawa*. Ten controls stated that current SRs should be replaced every 30 days or less. Four asked for replacement every two weeks. Controls generally asked for more frequent replacements because they reported that the active ingredient quickly became depleted. One control group participant suggested a reformulated SR with a replacement interval greater than 6 months; others favored SRs with sufficient *dawa* for 3 months.

Participants were also asked about installation preferences. Some who initially refused hooks to avoid having holes drilled in their walls later requested them since SRs installed with tape often fell. Tape also left adhesive marks on the walls, making the house unattractive (Figure 5).

**Figure 5:**
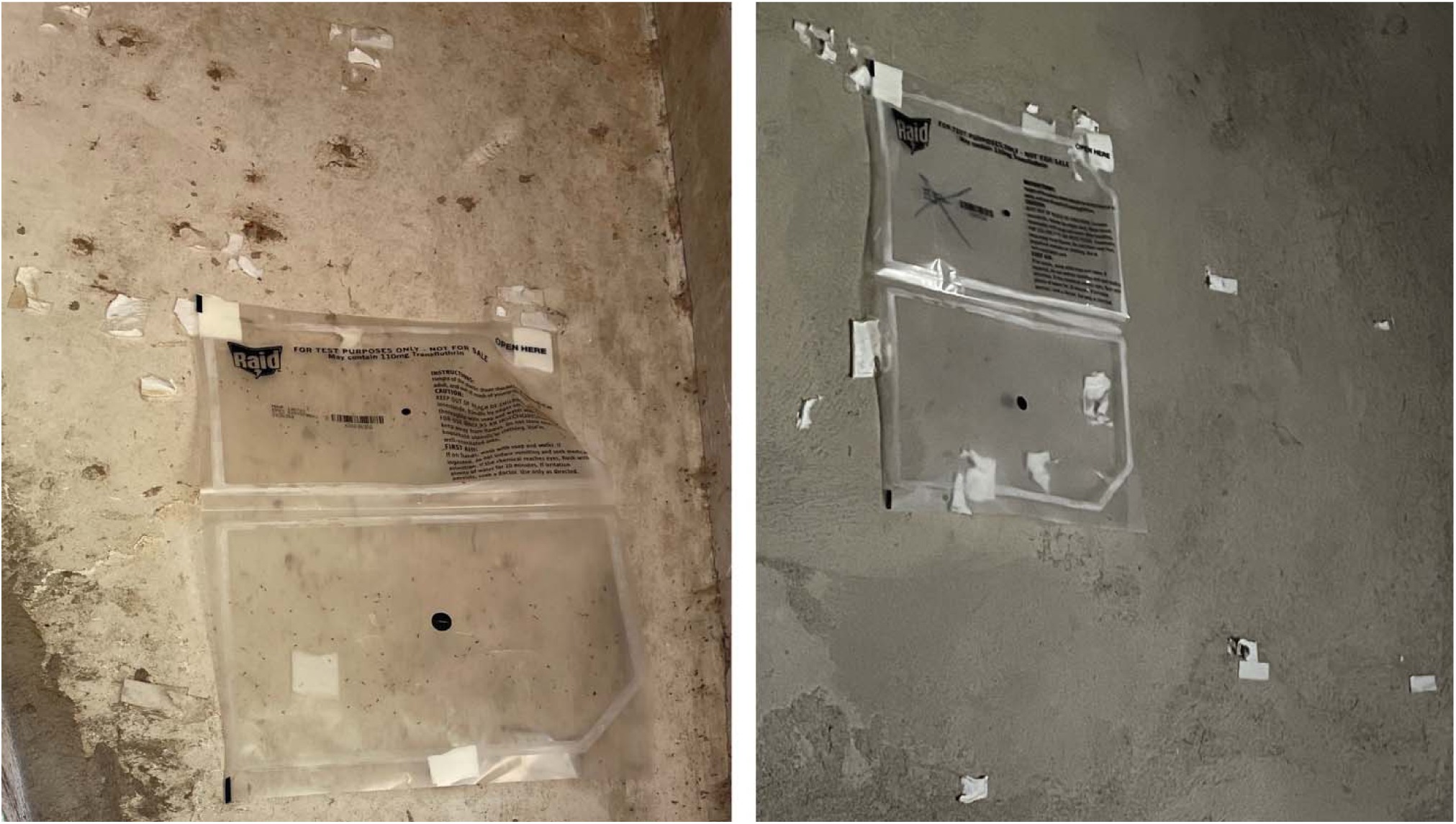
Adhesive marks left on walls from installation with double-sided tape.

## Discussion and Recommendations

### Participant experience with Mosquito Shield™ Perceived efficacy

TIPs intervention participants in this study perceived Mosquito Shield™ to be effective and acceptable. While some reported fewer cases of malaria among family members, most seemed to assess effectiveness based upon perceived mosquito density. This despite entomological findings that did not demonstrate a statistically significant decrease in mosquito density.[17] Participant focus on mosquito density may be associated with the presence of all mosquito species, not just malaria-transmitting anophelines. This is consistent with participant reports of less buzzing and less aggressive mosquito behavior. It is also consistent with decades of research showing that net users often value ITNs as much for their ability to reduce so-called “nuisance” biting (by non-malaria transmitting mosquitoes) as for their protection against malaria. [23–25]

Potential reasons why control participants reported early in the trial that their placebo products were working could include placebo effect, social courtesy or social desirability bias, or seasonal fluctuations in mosquito populations. The Busia County ITN distribution campaign shortly before study launch could also have contributed to perceived efficacy of the placebo product early on.

Several intervention and control participants described Mosquito Shield™ as more effective in some rounds and less effective in others, suggesting possible miscommunication about study design. While cRCT study team members informed enrolling participants that they would be assigned to either intervention or control but would not know which, it appears that some participants, some CHVs, or both interpreted this to mean that any given household might sometimes receive active SRs and sometimes placebos. In one participant’s words:

> You know, the people who come to install [the SRs] are saying sometimes they get the ones without ekeya. Sometimes they do bring the one that has, what? Ekeya. That is how they change. When you see the ones and the mosquitoes are there, that one has nothing. If you see the ones with one mosquito here and one mosquito there, those [are the] ones [that] have ekeya.” (Male, 50’s, control, round 4)However, product management monitoring indicated that no such crossover occurred.

### Acceptability

Most intervention participants remained consistent throughout the trial about their desire to keep using Mosquito Shield™ in their households. In contrast, controls reported growing doubts, citing inconsistent effectiveness and negative community feedback. By later product replacement rounds, many stated that they saw no reason to keep accepting the SR since they perceived it as offering no protection. Those who continued to welcome installation described the product as having some effect even if not as much as expected. Rather than outright rejection, these participants often called for stronger repellent (more *dawa*), reinforcing the link between acceptability and perceived efficacy. Factors favoring acceptance in both arms included the SR’s mosquito-repelling effects, its ease of use compared to ITNs, and its ability to protect people during waking hours when ITNs were not in use.

### Perceived side effects

Epidemiological data from this and previous SR trials shows no association with serious adverse effects (SAEs).[17, 26, 27] Nevertheless, user perceptions of side effects could significantly impact acceptability even if epidemiological data demonstrate that such perceptions are unwarranted.

Kenya TIPs intervention participants reported few perceived side effects from Mosquito Shield™ and described any they did perceive as tolerable given the SR’s benefits. Controls, who received placebos and thus should have experienced no actual SR-attributable side effects, reported perceiving them in a larger proportion of interviews (19%) than intervention participants (11%). This suggests that such effects were likely due to unrelated causes.

### Use of ITNs and other mosquito control products and practices

Despite repeated guidance to keep using ITNs because SR efficacy was still unclear and households would not know if they were receiving an active SR or a placebo, many participants reported discontinuing net use. Upon receiving such a report, social science team members shared the information with the cRCT coordinator who then arranged additional cluster-level behavior change communication reemphasizing the importance of continued use. This included in-person reminders from study CHVs as well as radio announcements in the study area. The overall study team agreed to this cluster-level strategy to avoid singling out a particular household for attention and perhaps compromising relations between that household and the study. Nevertheless, study participants continued to report only intermittent use. Future SR interventions should anticipate a need to address this issue. Conversely, participant reports of saving money by no longer needing to purchase mosquito coils may serve as a promotional point for SR adoption.[28] Decreased respiratory discomfort due to the absence of smoke from burning coils, leaves, or other combustibles used to repel mosquitoes may likewise increase SR appeal.

### Methodological innovation

This study’s link to a 24-month efficacy trial provided unique methodological opportunities. TIPs typically take place over a few weeks or months to pilot test a behavioral intervention.[29] Our 5 TIPs visits over 18 months offered insight into the evolution of participant response to Mosquito Shield™ from one week post-installation through 18 cycles of 28-day replacement. This helped us assess not just initial acceptance but change in perceived efficacy, acceptability, and factors affecting sustained adoption over time. Our work both draws from and contributes to the relatively limited literature on longitudinal qualitative analysis.[30]

Further, while TIPs were developed to test behavior change interventions, this study adds to previous literature confirming the value of TIPs for pilot-testing products and soliciting end-user improvement suggestions.[18, 31, 32] In this way, TIPs function as a variant of human-centered design for health promotion and disease prevention. Finally, the cRCT’s design provided an opportunity to conduct a double-blinded longitudinal social science study, something relatively rare in a field where most interventions are, by nature, not blindable.

### Added value to parent efficacy trial

This study illustrates the added value of nesting social science research within an efficacy trial. First, the results reported here enhance our understanding of end-user perspectives about the proposed intervention. TIPs data shed additional light on the built environment in which SRs would be used: housing type, construction materials, floor plans, living and sleeping spaces, and day-to-day practices (cooking, decorating, storage). It helped identify initially insignificant-seeming issues that might prove critical to SR acceptance such as user preference for installation with hooks rather than tape. It likewise revealed user preferences about product characteristics: While priorities may differ by site, Kenyan participants made clear that minimizing number of units required and maximizing time between replacement outweighed preferences for color, size, shape, or fragrance.

This study also helped us determine how SRs might affect current mosquito control products and practices. Impacts included some undesirable effects (reduced ITN use) and some neutral or potentially desirable ones (reduced use of mosquito coils, topical repellents, and home-based practices such as burning leaves). Such knowledge is useful whether SRs ultimately become a public- or private-sector intervention.

Second, concurrent social science research builds a foundation for more rapid scale-up. Our findings have identified factors (e.g., installation method) likely to affect sustained uptake. They have identified challenges (e.g., decreased ITN use) and generated some preliminary data on how to address them. Without such research during the cRCT, scale-up would likely take longer and prove more problematic. Identifying and trouble-shooting such issues on a small scale in advance is more efficient and less costly than doing so during mass implementation and may create fewer long-term barriers to adoption.

Nested social science research helped strengthen the parent cRCT, however, even before consideration of scale up or impact. Though a relatively small subset of the sample, TIPs households contributed to assuring cRCT adherence to protocol. When a social science team member observed any anomaly – delayed or non-standard SR installation for instance – the team member would ask the participant about it and report back to the cRCT. The cRCT would then determine an appropriate solution, incorporating participant input if appropriate. The social science team’s training in qualitative and ethnographic methods and its rapport-building with TIPs participants enabled team members to not only *identify* deviations but also establish their *root causes.* This helped the cRCT respond to deviations rapidly and thoroughly, assure a return to protocol adherence, and minimize potential threats to study validity or outcome. In all cases the objective of identifying and resolving deviations was not to find fault but to ensure the strongest possible trial outcome. The social science study thus helped increase the likelihood of a definitive cRCT outcome and decrease the threat of an indeterminate one.

### Limitations

While this study offers useful insights, it is limited by sample size and geographic coverage. The qualitative methods used here are designed to provide in-depth information about human behavior – in this case perceived efficacy and acceptability of SRs in the study area. Though transferable, the methods are not intended to produce generalizable conclusions. Similarly, the study’s purposive sampling is designed to identify participants capable of providing substantive feedback over time. This produces richer, more detailed data than a population-based random sample, but is not intended to be generalizable.

Reporting counts and percentages in qualitative studies is risky: Not all themes are discussed in all interviews and any given theme may be discussed differently with different participants or in different interviews with the same participant. Thus, percentages are not necessarily based on a common denominator. Here we reported the percentage of interviews in which participants ruled out, expressed uncertainty about, or reported perceived SR-related side effects. Many more participants denied side effects than reported them, but this should not be interpreted as a definitive measure.

This study did not address distribution methods for Mosquito Shield^TM^ in a non-research setting, nor whether end users are capable of installing the product safely and effectively, replacing it at prescribed intervals, or disposing of it safely. Safe disposal is a particular concern since Mosquito Shield™ is plastic and requires disposal that minimizes environmental harms. Moreover, even depleted SRs retain and continue to disperse residual transfluthrin which is toxic to fish and other aquatic organisms.[33]

During the trial, study CHVs retrieved depleted units while installing replacements, then surrendered them to study staff for high-temperature incineration. Since coordinated disposal requires systematic retrieval and since high-temperature incinerators are not widely accessible, the study disposal method is unlikely to function effectively at scale. Additional operations research will be needed to address these issues.

### Recommendations

Future studies and implementation programs should keep in mind the following recommendations:

1. **Emphasize that SR recipients should continue to use ITNs and accept IRS.** More thorough community dialogue should highlight the fact that SRs are meant to complement rather than replace other mosquito control products and practices, thus expanding the time and physical space in which household members are protected. Results from this study showed that many study participants stopped using ITNs if they perceived the SRs to be effective.
2. **Develop a longer-lasting SR capable of protecting more space.** Kenyan study participants stated that the 28-day replacement interval for Mosquito Shield™ was too short. They also found installing so many units unsightly. A longer-lasting SR capable of protecting more space will likely prove more acceptable.
3. **Pilot test SR installation methods.** Installation methods affect both deployment cost and acceptability. If SRs are installed using an objectionable method, users are more likely to reject them. Kenyan users preferred hooks to tape and commented that tape left ugly adhesive marks on their walls. Installation preferences may vary by setting, making site-based pilot testing critical.
4. **Develop a biodegradable product medium.** Biodegradable SRs would be preferable to plastic. Some Kenyan participants expressed concern about the environmental impact of discarding used plastic sheets, especially given the current 28-day replacement cycle and the need for many units.
5. **Address potential safety concerns.** While perceived side effects were few, some intervention and control participants did raise concerns about human and environmental safety. Failure to acknowledge and address such concerns adequately – even if epidemiological surveillance data show no evidence of adverse events – can lead to rejection of even very safe public health interventions and distrust of public health authorities.[34] Community entry and dialogue should explore local safety concerns and share safety data in a manner accessible to the local population. Further research on how best to summarize and present safety data may aid SR adoption. Participant reports in the present trial that the product was odorless and did not cause respiratory problems for asthmatics may help allay potential concerns about respiratory issues in future implementation. Future efforts should consider including this finding among supportive health communication messages.
6. **Test SR use in community settings.** Future research should explore SR efficacy and acceptability in public spaces such as schools, public markets, private businesses, and houses of worship. National vector-borne disease control programs, implementing partners, and donor agencies should consider potential SR use in such spaces to complement household vector control.

## CRediT authorship contribution statement

**Steven Harvey:** Writing-original draft, Writing-review & editing, Methodology, Formal analysis, Supervision, Project Administration, Visualization, Resources, Validation, Conceptualization Funding acquisition. **Sheila Muya Ekodir:** Validation, Formal Analysis, Data Curation, Writing-review & editing, Investigation. **Lucy Baker:** Writing-review & editing, Data curation, Formal analysis, Project Administration, Validation, Investigation. **Julius Ichodo Odero:** Investigation, Writing-review & editing, Formal Analysis, Validation. **Moureen Ekisa:** Investigation, Formal Analysis, Validation. Writing-review & editing, **Jane Ikapesi:** Investigation, Formal Analysis, Validation. Writing-review & editing, **Samantha W. Tsang:** Methodology, Formal Analysis, Validation, Writing-review & editing. **Albert Casella:** Validation, Writing-review & editing, Formal Analysis. **Kaci McCoy:** Data Curation, Project Administration, Writing-review & editing. **Anna Passaniti:** Validation, Writing-review & editing. **Quentin Awori:** Investigation, Writing-review & editing. **Bernard Abong’o:** Investigation, Writing-review & editing. **Brian Polo:** Investigation, Writing-review & editing. **Vincent Moshi:** Data Curation, Validation, Formal Analysis, Writing-review & editing. **Sean Blaufuss:** Validation, Writing-review & editing. **Manothri Mallikarachchi:** Validation, Writing-review & editing. **Ashley Hudson:** Project Administration, Supervision, Writing-review & editing. **Nicole L. Achee:** Funding acquisition, Supervision, Writing-review & editing, Conceptualization. **John P. Grieco:** Funding acquisition, Supervision, Conceptualization. **Eric Ochomo:** Writing-review & editing, Supervision, Conceptualization.

## Supporting information

WHO ERC approval

## Data Availability

All data produced in the present study are available upon reasonable request to the authors

## Acknowledgements

We are especially indebted to the residents of Teso North and Teso South Districts; we are grateful for their participation and their willingness to have this study be conducted in their communities. We would also like to thank the community leaders and health promoters for recruiting participants. We would like to thank the Busia County Health Department for facilitating the study in Busia. We are grateful to the KEMRI leadership and staff for their institutional support, institutional review board guidance, and overall support. We would like to thank Evercita Eugenio for her support as the external statistician on the data safety and monitoring board (DSMB). We would also like to thank SC Johnson for their industry and product expertise as well as their independent funding of the development, manufacturing, delivery, and shipment of the spatial repellent used in this study.

