## Supplementary material for "Perceived Efficacy and Acceptability of a Spatial Repellent for Malaria Control in Western Kenya: Lessons from a Qualitative Social Science Study Conducted in Conjunction with an Epidemiological Efficacy Trial": WHO ERC approval

***WHO ERC***
**Amendments**

**Protocol ID**: ERC.0003186 UND-SocialScience (Kenya)
**Protocol Title:** Understanding behavioral aspects of household spatial repellent use for malaria control in western Kenya
**Version: --- Dated:** 17/08/2019
**WHO Responsible Staff Member**: Ekaterina RYKOVANOVA
**Responsible Unit**: Unitaid

Dear Dr. Ekaterina RYKOVANOVA,
Thank you for your submission requesting approval of amendments. This protocol received original approved by the ERC on 06.09.2019. The outcome of review is as follows:

1. It is noted that the ethics committee from the Kenya Medical Research Institute (KEMRI) requested the addition of a witness signature to all consent forms and they were modified following this request.
2. No changes to the WHO ERC approved protocol (Protocol dated 17.08.2019) were made.
3. The data collection instrument was modified to reflect the information obtained during pre-testing.

The following documents are approved as submitted:

Consent Forms (English version):
JHU-SS-CF-A-B-FreelistingRanking_v3.0_20191014_Track Changes
JHU-SS-CF-A-B-FreelistingRanking_v3.0_20191014_Clean Copy.docx
JHU-SS-CF-C-IDI_v3.0_20191014_Clean Copy.docx
JHU-SS-CF-C-IDI_v3.0_20191014_Track Changes
JHU-SS-CF-D-HouseholdObs_v3.0_20191014_Clean Copy.docx
JHU-SS-CF-D-HouseholdObs_v3.0_20191014_Track Changes
JHU-SS-CF-E-TIPs_v3.0_20191014_Clean Copy.docx
JHU-SS-CF-E-TIPs_v3.0_20191014_Track Changes
JHU-SS-CF-F-RetailAudit_v3.0_20191014_Clean Copy.docx
JHU-SS-CF-F-RetailAudit_v3.0_20191014_Track Changes

Data Collection Instruments (English version)
JHU-SS-DCI-A-Freelisting_v2.0 _20191024_Clean Copy.docx
JHU-SS-DCI-A-Freelisting_v2.0_20191024_Track Changes.docx
JHU-SS-DCI-B1-Rating_v2.0_20191017_Clean Copy.doc
JHU-SS-DCI-B2-Ranking_v2.0_20191017_Clean Copy.doc
JHU-SS-DCI-B2-Ranking_v2.0_20191017_Track Changes
JHU-SS-DCI-B2-Rating_v2.0_20191017_Track Changes JHU-SS-DCI-C-IDI_v1.0_20190412.docx
JHU-SS-DCI-D- HouseholdObs_v1.0_20190412 JHU-SS-DCI-E-TIPs_v1.0_20190415.docx
JHU-SS-DCI-F-RetailAudit_v3.0_20191024_Clean Copy.docx
JHU-SS-DCI-F-RetailAudit_v3.0_20191024_Track Changes
JHU-SS-DCI-G-KII-v0.2 20190414

**Chairperson …………………………………….** **Date.** 11.11.2019

**Name: Peter Olumese**
